# Fasting Plasma Glucose as a Primary Screening Test for the Diagnosis of Gestational Diabetes mellitus

**DOI:** 10.1101/2024.08.23.24312519

**Authors:** Md. Rakibul Hasan, Nusrat Sultana, Sandesh Panthi, Mashfiqul Hasan, Sharmin Jahan, Yasmin Aktar, Muhammad Abul Hasanat

## Abstract

**Aim:** To see the diagnostic efficacy of fasting plasma glucose (FPG) in respect to OGTT during pregnancy.

**Materials and Method:** In this cross-sectional study, we enrolled 542 pregnant women 18 years or older by consecutive sampling irrespective of gestational age. 75gm three samples OGTT was done and categorized them to either normal glucose tolerance (NGT) or abnormal glucose tolerance (AGT) according to World Health Organization (WHO) 2013 criteria.

**Results:** The sensitivity of FPG with a threshold of 5.1mmol/L was 47.3% among the study subjects, which was very low but specificity was very high (99.7%). However, changing the cut off value to 4.7mmo/L and 4.5mmol/L significantly increased the sensitivity to 64.8% and 73.3% with modestly decreased specificity to 86.2% and 73.2% respectively. In pregnant women with gestational age 24 to 28 weeks, FPG with a threshold of 4.5mmol/L could identify 75% of GDM subjects with specificity 76.8%. Furthermore, it has the highest sensitivity in detecting AGT in all three trimesters (80%, 74.4% and 68%) compared to 4.7mmol/L (80%, 68.9% and 50%) and 5.1 (80%, 47.8% and 30%) in 1^st^, 2^nd^ and 3^rd^ trimester respectively. FPG had positive correlation with maternal age (r=0.138, p=0.001), BMI (r=0.164, p<0.001) but negative correlation with weeks of gestation (r= -0.242, p<0.001).

**Conclusion:** FPG with cut off value of 4.5mmol/L may be used as an initial screening test for GDM to reduce the need for OGTT.

## Introduction

Gestational diabetes mellitus (GDM) is one of the most typically encountered endocrine issues in pregnancy. About one in six women globally and one in four women in Southeast Asia has been suffering from some form of hyperglycemia in pregnancy (HIP), and GDM contributes about 80% of cases of HIP.^1^ It is associated with many complications for both mother and offspring reported from many well-established studies from different parts of the world.^2-6^ Screening for GDM is essential as a high prevalence of GDM is observed in Southeast Asia. Although screening for GDM with an oral glucose tolerance test is a gold standard and ideal procedure, but it is inconvenient, unpleasant, time-consuming, and unsuitable for large-scale screening. Furthermore, oral glucose load is not well accepted by many patients. One study reported that nearly half of the pregnant women were extremely bothered with having to drink the glucose solution.^7^Conversely, fasting plasma glucose (FPG) is inexpensive, reproducible, and suitable for a screening test. In the Hyperglycemia and pregnancy outcome (HAPO) study, 51% of GDM was diagnosed by an FPG > 5.1 mmol/L.^8^In China, Due to complexity of Chinese vast geography and population, they have adapted a two steps strategy for GDM screening with FPG. They selectively do OGTT in pregnancy if FPG value is found between 4.4 to 5.0mmol/L. This simplicity of test procedure ensures universal screening of GDM in low resource setting. ^9^In one study from South Africa reported, FPG of ≥5.1 mmol/L identified 87.8% of GDM patients and the sensitivity and specificity of FPG ≥4.5 mmol/L were 98% and 80%, respectively.^10^Our study aimed to see the sensitivity and specificity of FPG with different cut-off values as a screening test for GDM in different trimesters.

## Materials & method

### Study design & Study subjects

This cross-sectional observational study included 542 pregnant women with an age ≥18 years. Pregnant women with gestational age ≥ 6 to 37 weeks, irrespective of presence or absence of risk factors for GDM were recruited between February 2014 and January 2017 by consecutive sampling from the Antenatal clinic, Department of Gynecology & Obstetrics, Bangabandhu Sheikh Mujib Medical University (BSMMU), Dhaka, after approval by the Institutional Review Board (IRB) of the same institution. OGTT was done at the GDM clinic, Department of Endocrinology, BSMMU. Any women with prior history of GDM or DM, or currently using steroids at supraphysiological dose for any illnesses were excluded from the study. Three-sample 75-gm OGTT was done and categorized them into either NGT or AGT according to WHO 2013 criteria. AGT comprises both GDM and DIP. Demographic and clinical variables including height, weight, BMI (kg/m^2^), and blood pressure (mm Hg) were recorded in structured data collection sheet for analysis.

### Analytic method

After collection of blood samples, it was centrifuged at the collection site within 15 minutes and transported to biochemistry laboratory within one hour of collection for analysis. Plasma glucose was assayed by the glucose oxidase method with an automated analyzer [RA-50 analyzer (Dade Behring, Germany)].

### Statistical analysis

All data were processed in IBM SPSS Statistics for Windows version 23.0 (IBM Corp, Armonk, NY, USA) and expressed as frequencies or percentages as well as mean (± SD) as applicable. The frequency of AGT was assessed by descriptive statistics and comparison of different variable between NGT and AGT were done by using chi-square test and independent sample t-test as applicable. The p values ≤ 0.05 were considered statistically significant. Correlation of FPG with various maternal factors was done by Pearson correlation. Sensitivity, specificity, positive predictive value and negative predictive value of FPG were assessed inrelation to three samples OGTT status according WHO 2013 criteria using following formulas.

**Formula 1:** Accuracy of a diagnostic test (FPG)

| **Diagnostic Test (FPG)** | **WHO 2013 Criteria** |  |
| --- | --- | --- |
|  | **Affected** | **Unaffected** |
| **Positive test** | True positive (a) | False positive (b) |
| **Negative test** | False negative (c) | True Negative (d) |

## Result

Our study populations were mainly from housewife occupational category (69%) and majority of them was multigravida (59%). About half of the study populations (49.1%) were from second trimester and only 16% of them from first trimester (Table 1). Based on WHO 2013 criteria, 30.5% of the study populations had AGT, of which GDM were 26.4% and DIP were 4.1% as depicted in figure 1. The sensitivity of FPG as a screening test for GDM compared to three samples 75g OGTT was 47.5% among the study subjects, which was very low but it’s specificity, positive predictive value, negative predictive value were seemed to be good (99.73%, 98.73% and 81.21% respectively). However, changing the cut off value of FPG from 5.1mmol to 4.7mmo/L and 4.5mmol/L significantly increased the sensitivity to 64.8% and 73.3% respectively (Figure 2). Selecting the study population between 24-28 weeks of gestation mildly increased the diagnostic sensitivity and specificity of FPG and a threshold of 4.5mmol/L could identify 75% of GDM subjects with specificity 76.8% (Figure 3). Trimester specific analysis of efficacy of FPG revealed the FPG has the highest sensitivity for diagnosis of GDM in first trimester (80%) whatever the cut-off value used for diagnosis, but its sensitivity progressively declined in subsequent trimesters (Table 2). Among different cut off value of FPG, 4.5mmol/L had the highest sensitivity in all three trimester for diagnosis of GDM (Table 2). Pearson correlation revealed FPG had positive and significant correlation with maternal age and BMI; however it was negatively correlated with gestational weeks (Table 3).

**Table 1:**
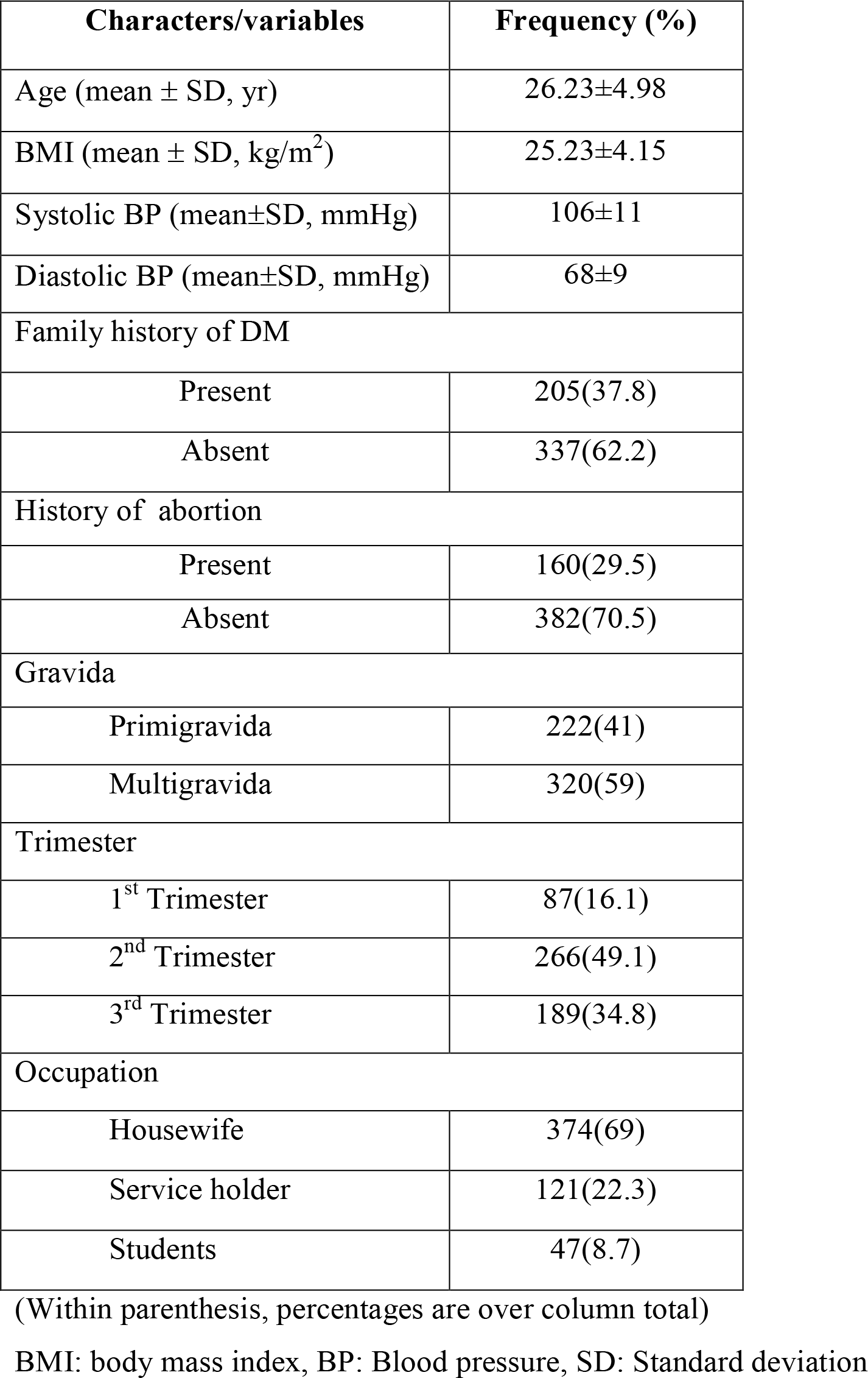
Characteristics of study subjects (N=542)

| Characters/variables | Frequency (%) |
| --- | --- |
| Age (mean ± SD, yr) | 26.23±4.98 |
| BMI (mean ± SD, kg/m<sup>2</sup>) | 25.23±4.15 |
| Systolic BP (mean±SD, mmHg) | 106±11 |
| Diastolic BP (mean±SD, mmHg) | 68±9 |
| Family history of DM |  |
| Present | 205(37.8) |
| Absent | 337(62.2) |
| History of abortion |  |
| Present | 160(29.5) |
| Absent | 382(70.5) |
| Gravida |  |
| Primigravida | 222(41) |
| Multigravida | 320(59) |
| Trimester |  |
| 1<sup>st</sup> Trimester | 87(16.1) |
| 2<sup>nd</sup> Trimester | 266(49.1) |
| 3<sup>rd</sup> Trimester | 189(34.8) |
| Occupation |  |
| Housewife | 374(69) |
| Service holder | 121(22.3) |
| Students | 47(8.7) |

(Within parenthesis, percentages are over column total)
BMI: body mass index, BP: Blood pressure, SD: Standard deviation

**Table 2:**
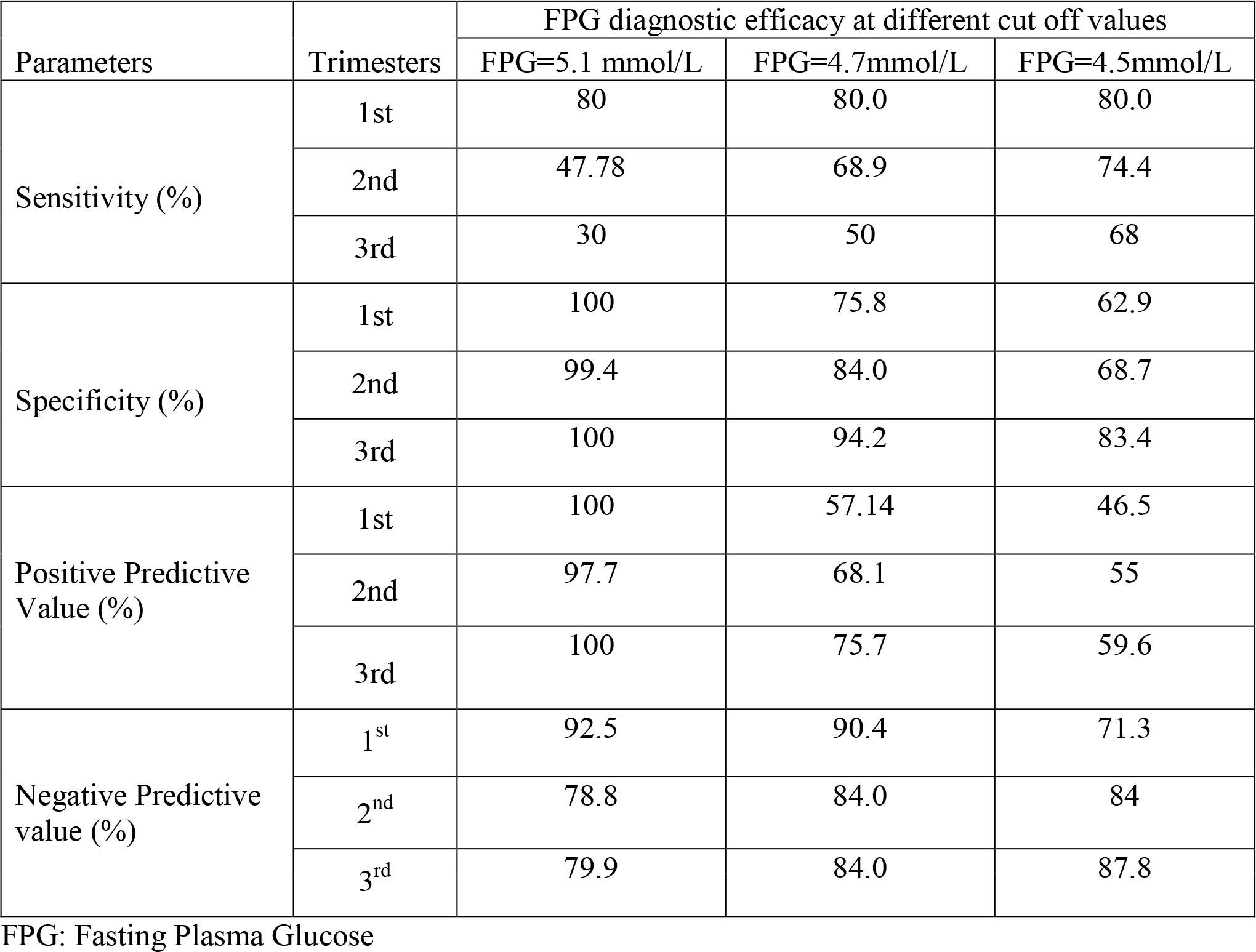
Diagnostic efficacy of Fasting Plasma Glucose with cut off value of 5.1, 4.7 and 4.5 mmol/L for diagnosis of Abnormal Glucose Tolerance at different trimester (N=542)

**Table 3:** Pearson correlations of FPG with various patient factors.

| Determinant | Study Subjects (n=542 ) |  |
| --- | --- | --- |
|  | r | p |
| FPG vs. Age | 0.138 | 0.001 |
| FPG vs. BMI | 0.164 | <0.001 |
| FPG vs. Weeks of gestation | -0.242 | <0.001 |

FPG: Fasting plasma glucose, BMI= Body mass index ( $\text{kg/m}^2$ )

**Figure 1:**
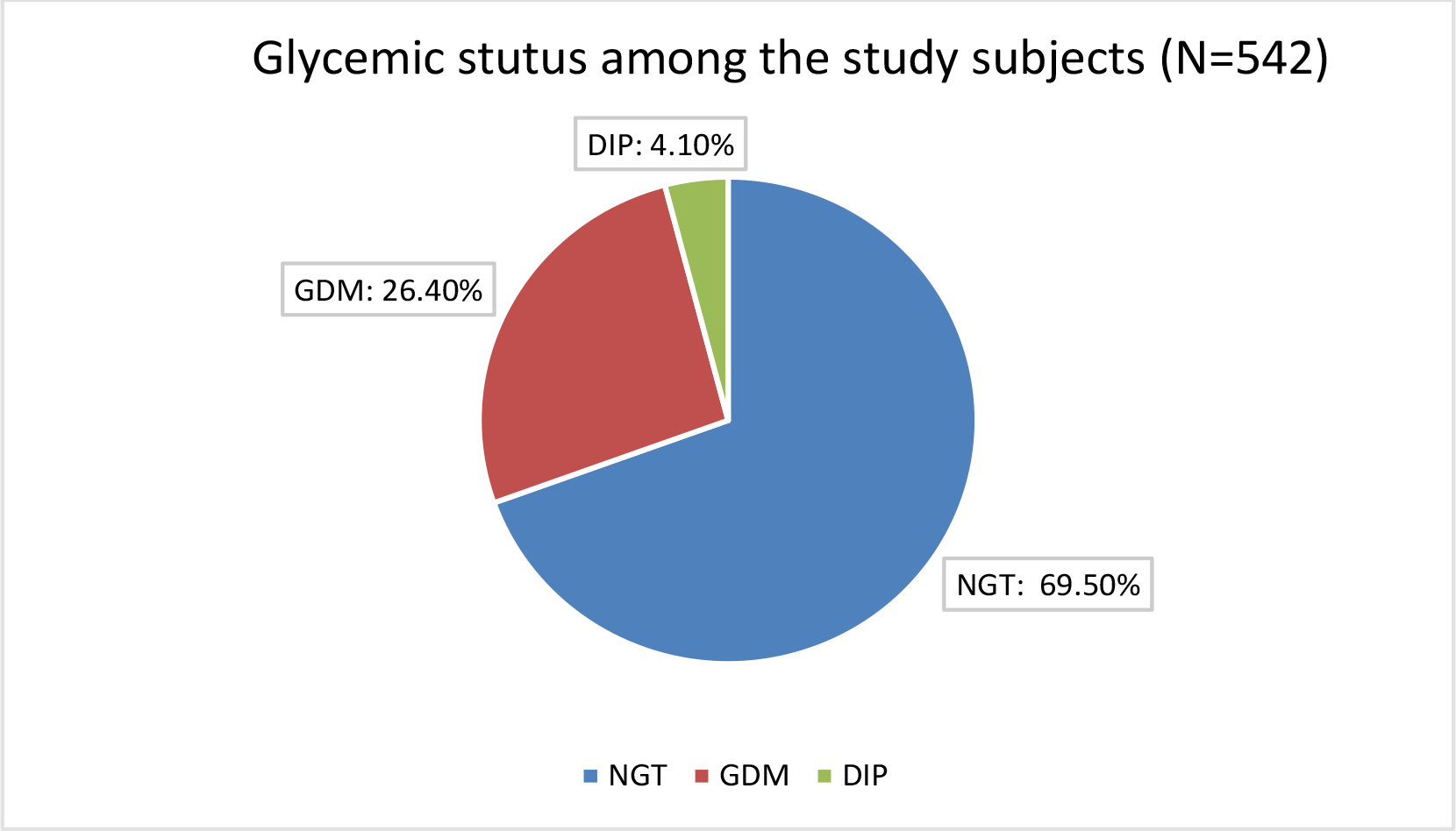
Glycemic status among the study subjectsaccording to WHO 2013 criteria. (NGT= Normal Glucose Tolerance, DIP= Diabetes in Pregnancy, GDM= Gestational Diabetes mellitus)

**Figure 2:**
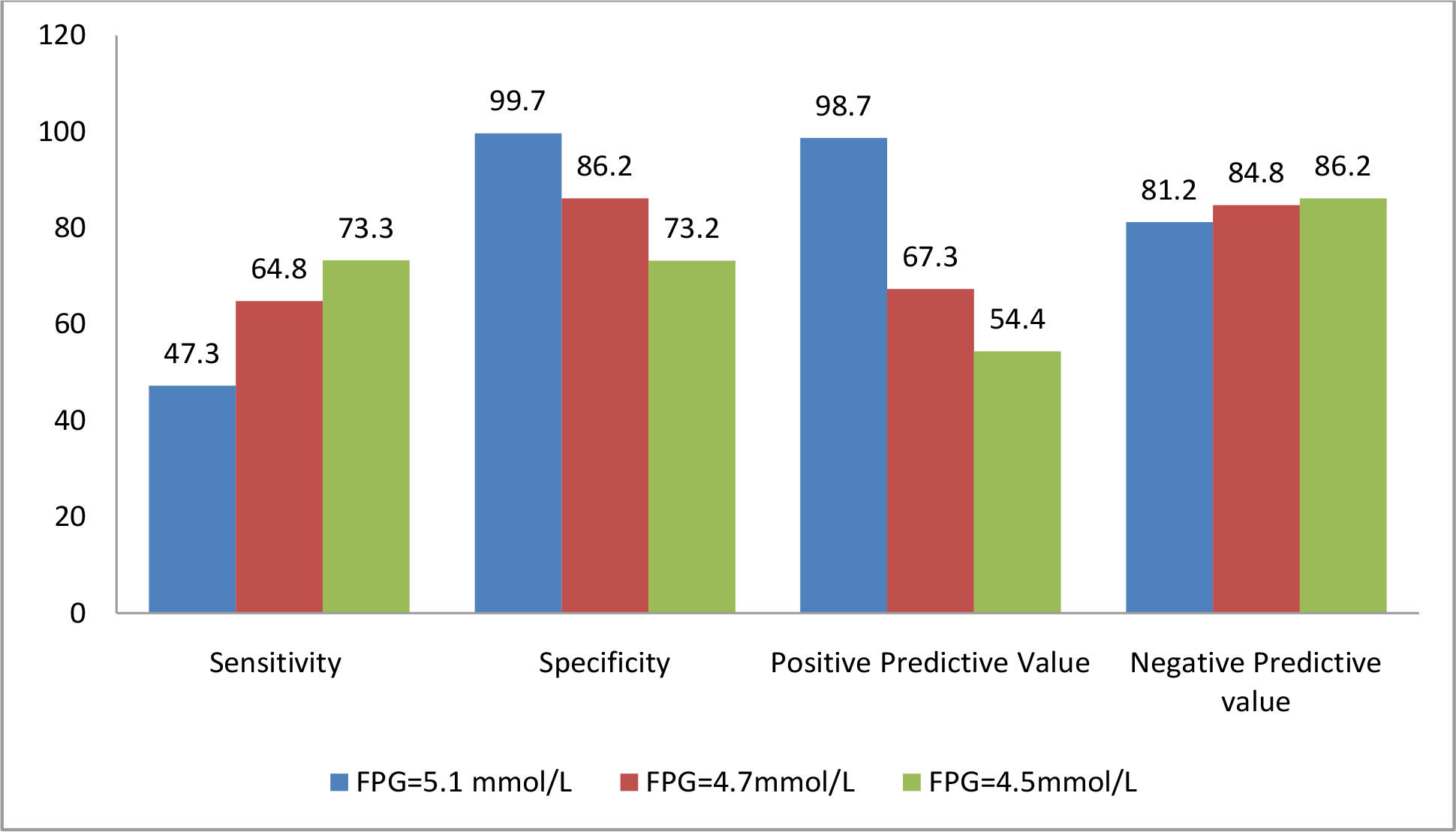
Diagnostic efficacy of Fasting Plasma Glucose in study subjects of all trimester (n=542) with different cut off value (5.1, 4.7 and 4.5 mmol/L)

**Figure 3:**
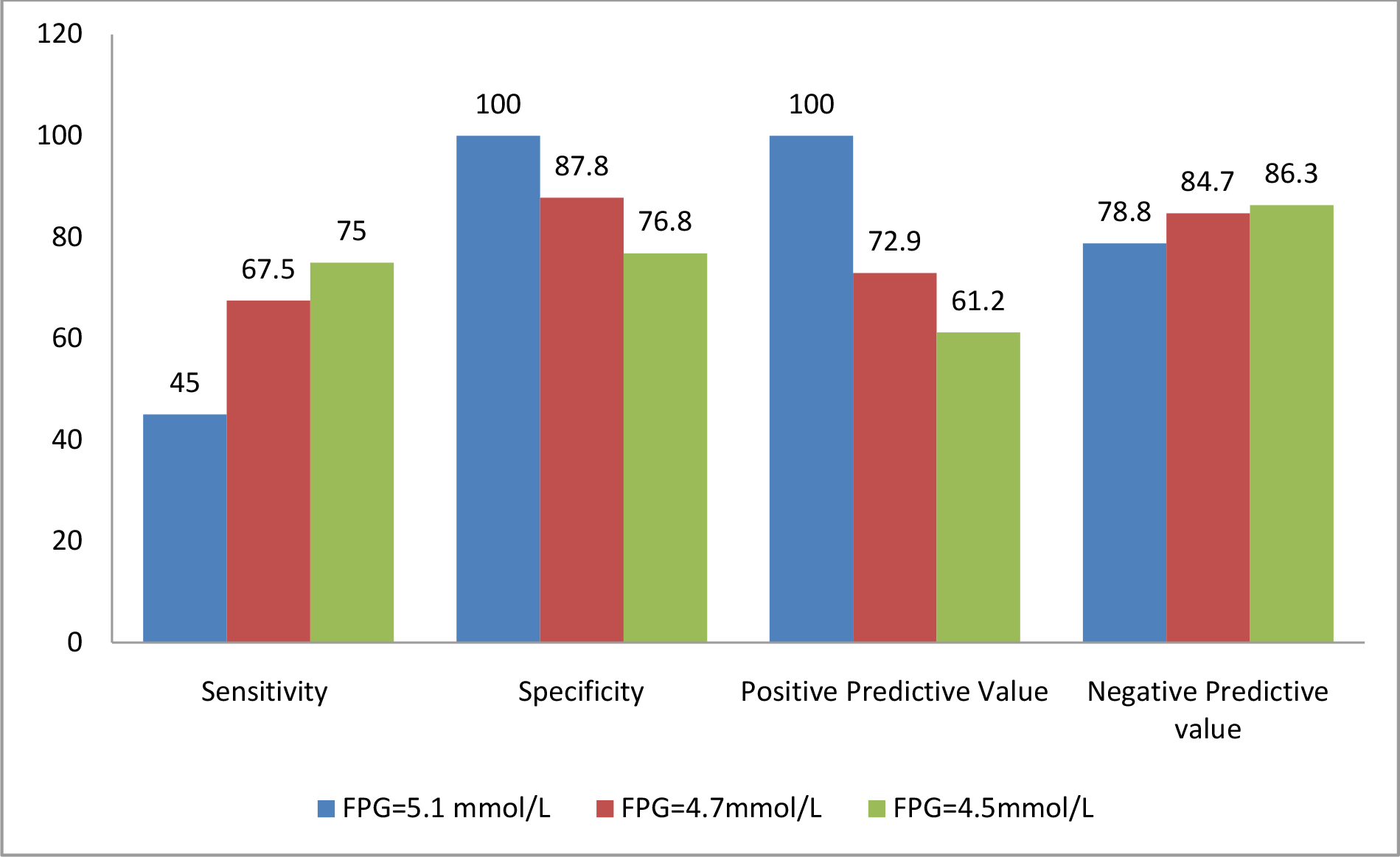
Diagnostic efficacy of Fasting Plasma Glucose in pregnant women with gestational age 24-28 weeks (n=122) with different cut off value (5.1, 4.7 and 4.5 mmol/L)

Figure-4 shows the receiver operating characteristic (ROC) curve showing the performance of FPG in diagnosing AGT of pregnancy (AUC 0.80; 95% CI: 0.76-0.85; p<0.001). The Youden index was 0.56 at the FPG cut-off 4.59 mmol/L with a sensitivity of 70% and specificity 81%.

**Figure-4:**
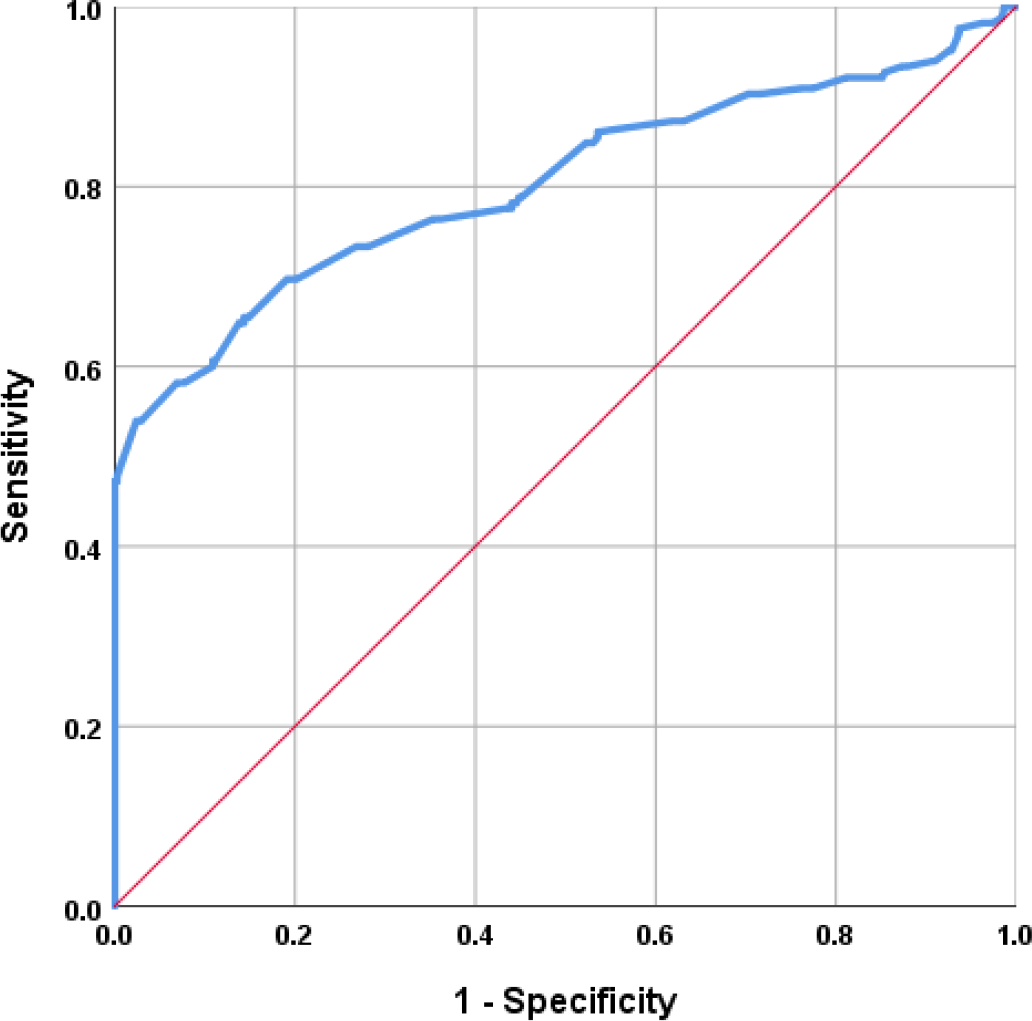
The Receiver Operating Characteristic (ROC) curve showing the performance of FPG in diagnosing AGT in pregnancy among the study subjects (N=542) FPG: fasting plasma glucose, AGT: abnormal glucose tolerance (includes gestational diabetes mellitus and diabetes in pregnancy)

## Discussion

It is well established from pregnancy outcome based study that the OGTT is a gold standard for diagnosis of GDM.^11^ However many pregnant women feel distressed during oral glucose load for OGTT.^7^ The use of FPG as an screening test to avoid OGTT was studied by many researcher in different country with very positive result.^10,12-16^ In our study, FPG could identify only 47.5% pregnant women with GDM with the same cut off value used in WHO 2013 criteria and international association of diabetes in pregnancy study group (IADPSG) criteria, which is a little bit similar to a study done by Agarwal MM et al. ^17^ However, changing the cut off value of FPG from 5.1mmol to 4.5mmol/L significantly increased the sensitivity to 73.3% and specificity 73.2%. A study done among the Mexican population reported that the FPG threshold of 4.5 mmol/L had the higher sensitivity (97%) but the specificity was very poor (50%). ^13^This significant difference in sensitivity and specificity may be attributed by characteristics of study population. Mean age of our study population was 26 years, whereas it was 16 years and only included adolescent women in that study. Maternal age has significant impact on overall prevalence of GDM as well as the FPG during pregnancy, Study reported that the addition of maternal age to FPG increase the sensitivity of FPG for diagnosis of GDM. ^14^Our study also reported significant positive correlation of maternal age with fasting plasma glucose.

Osman Şevketet. al reported the higher sensitivity (93.4%) of FPG cut-off value of ≤4.5 mmol/l with almost similar specificity (67.6%) like our study.^15^ A study from South Africa reported very high sensitivity and specificity (98% and 80% respectively) with the same FPG cut off value,^10^ this may be due to gestational age of the study population, they included only the pregnant women with gestation age 24-28 weeks whereas our study population were from all three trimesters. Even though, subgroup analysis with pregnant women of 24-28weeks of gestation in our study population, revealed relatively lower sensitivity and specificity. Ethnicity may also contribute to the sensitivity which needs to be explored by multinational study including the population of different ethnicity.

Since 2000, the UAE has been using two thresholds for “rule-in and rule-out” of GDM based on multiple studies which significantly reduce the number of OGTT. They used FPG cut-off of 5.0mmol/L for “rules-in” GDM with 100% specificity and another cut-off of 4.4mmol/L for “rules-out” GDM with variable sensitivity. Using this approach successfully avoided up to 70% OGTT during pregnancy. Similar results were observed from studies in China and Brazil also.

^18^Our study also observed similar result; using FPG with a threshold of 4.5mmol/L could potentially identify about 70% pregnant women with GDM and reduce the need for oral glucose load in a significant number of patients.

### Limitation

Pregnant women in all trimesters were included in our study, whereas most of the studies included only pregnant women with gestational age 24 to 28 weeks, using IADPSG or WHO criteria. Relatively lower number of study population of this specific gestational week’s category may attribute to relatively lower sensitivity and specificity of FPG as a screening test in our study. Asian population typically has lower FPG with sharp rise of post prandial plasma glucose which is not usually seen in Caucasian counterparts. ^19^ So sensitivity of FPG may vary in different regions of the world.

## Conclusion

FPG may be used as a screening test for selecting the ideal patient for OGTT.Using a protocol adapted in the UAE and China may be a suitable alternative and cost effective strategy for universal screening of GDM.

## Data Availability

All data produced in the present study are available upon reasonable request to the authors

## Acknowledgements

We are grateful to the study participants and their attendants.

## Declaration

### Data Availability

Any inquiries regarding supporting data availability of this study should be directed to the corresponding author and are available from the corresponding author on reasonable request.

### Ethical approval

This study was approved by the Institutional Review Board (IRB) of BSMMU, Reg: No. BSMMU/2013/14269 and BSMMU/2016/2746.All procedures performed in studies involving human participants were in accordance with the ethical standards of the IRB and with the 1964 Helsinki Declaration and its later amendments or comparable ethical standards.

### Informed consent

Informed written consent was obtained from each of the participants included in the study.

